# Clinical outcomes of bio-interventional cyclodialysis with one or multiple allogeneic reinforcements: a multicenter real-world evidence study

**DOI:** 10.64898/2026.11.12.26367929

**Authors:** Craig Chaya, Nathan M Radcliffe, Mary Qiu, Sandra Sieminski, Shamil Patel, Tsontcho Ianchulev, Gabriel Ferreira, Manjool Shah, Jason Bacharach

**Author notes:** Corresponding Author: Tsontcho Ianchulev, MD, MPH, New York Eye and Ear Infirmary of Mount Sinai, 310 East 14th Street, New York, NY 10003, USA.

## Abstract

**Introduction:** Bio-reinforced cyclodialysis uses a biocompatible acellular scleral allograft spacer to maintain a surgically created cleft and enhance uveoscleral outflow. The titratable effect of 1 vs 2 spacers on intraocular pressure (IOP) lowering has not previously been evaluated.

Methods: Analysis of 260 eyes (187 patients) in the prospective, multicenter CREST study (NCT05506423) undergoing combined phacoemulsification and bio-reinforced cyclodialysis with one spacer (N = 146) or two (N = 114). The primary endpoint was IOP change from baseline to month 12. Analyses comprised unadjusted comparison, baseline-adjusted regression, 1:1 propensity-score matching, doubly robust overlap weighting, and models accounting for enrolling site and fellow eyes. IOP after a secondary IOP-lowering procedure was censored. IOP reported as mean (95% CI) and medications as mean (±SD).

Results: At baseline, IOP was 17.8 (4.2) versus 18.5 (5.7) mmHg and medications 1.3 (1.1) versus 1.5 (1.4), with one versus two spacers, respectively. Enrolling site was most imbalanced: one center contributed 38.6% of two-spacer eyes but 7.5% of one-spacer eyes. The unadjusted difference in IOP reduction was 2.5 mmHg (1.2–3.8; p<0.001) and persisted after propensity-score matching (2.4 mmHg; 0.9–3.8; p=0.001) and overlap weighting (2.0 mmHg; 1.2–2.8; p<0.001). After accounting for site, it attenuated to 0.9 mmHg (−0.1 to 1.8; p=0.080), and to 0.4 mmHg (−0.6 to 1.5; p=0.424) using both strategies. Medication reduction (−0.7 versus −0.6; p=0.643), freedom from secondary incisional surgery (98.6% versus 99.1%), and site-adjusted adverse-event rates did not differ.

Conclusion: Two-spacer treatment was associated with numerically greater 12-month IOP reduction in unadjusted and patient-level adjusted analyses, but this association was substantially attenuated after accounting for enrolling site. These short-term real-world data are compatible with, but do not establish, an independent incremental effect of a second spacer. Further study is warranted. Trial registration: ClinicalTrials.gov identifier NCT05506423.

**Plain Language Summary:** Glaucoma damages the nerve at the back of the eye, usually because pressure inside the eye is too high. One way to lower that pressure is an operation that opens a small channel so fluid can drain out of the eye more easily. A thin piece of donated, treated tissue from the white of the eye is placed there to hold it open. Surgeons can place one of these spacers or two. Nobody knew whether two lowers the pressure more than one. We studied 260 eyes treated at nine centers, all of which also had cataract surgery at the same time. Of these, 146 received one spacer and 114 received two. We compared how far the pressure inside the eye fell over the following year. Eyes that received two spacers had a larger fall in pressure, about 2.5 millimeters of mercury more than eyes that received one. That is the unit used to measure eye pressure. However, the surgeons who chose two spacers worked mainly at a few particular centers, and those centers differed from the others in many ways. When we compared eyes treated at the same center, the difference between one spacer and two almost disappeared. These findings suggest that two spacers may be associated with greater pressure lowering, but this study cannot determine whether the second spacer itself caused the additional effect. A randomized study, in which patients are assigned to receive one or two spacers, would be needed to answer that question more definitively.

**Key Summary Points Why carry out this study?:** • Bio-reinforced cyclodialysis maintains a surgically created cleft with an acellular scleral allograft spacer, and unlike most angle-based glaucoma surgery, in which the intervention is fixed, the number of spacers is a parameter the surgeon can titrate intraoperatively.

• We asked whether eyes receiving two spacers achieve greater 12-month IOP reduction than eyes receiving one, and whether any difference survives adjustment for measured baseline differences between the groups.

**What was learned from the study?:** • Two-spacer eyes had a 2.5 mmHg greater unadjusted reduction at 12 months, which persisted after propensity-score matching and overlap weighting but fell to 0.9 mmHg after accounting for enrolling site, to 0.4 mmHg within the sites that used both strategies, and was unchanged when fellow eyes were accounted for.

• Responder rates were numerically higher with two spacers but were attenuated after site adjustment, and adverse-event and secondary incisional surgery rates did not differ between groups; the study was not powered to establish safety equivalence.

Outcomes stratified by baseline visual-field severity showed no concentration of effect in more severe eyes, but the 139 eyes with baseline visual fields largely excluded the center contributing most two-spacer eyes, so the stratified analysis is effectively a within-center comparison rather than an independent test of effect modification.

Spacer number was strongly confounded by enrolling site, so a prospective randomized comparison is needed to determine whether a second spacer provides independent incremental IOP lowering, and observational analyses of surgeon-titrated interventions should account for both center- and patient-level factors.

## Introduction

The evolution of glaucoma surgery toward precision medicine demands interventions that can be tailored to individual patient needs. The American Academy of Ophthalmology Preferred Practice Pattern emphasizes that target intraocular pressure (IOP) should be individualized and may need adjustment during the course of disease [1]. However, most angle-based surgical devices offer a binary intervention with limited ability to titrate the magnitude of effect intraoperatively. This constrains personalized surgical planning across the heterogeneous range of glaucoma severity and IOP targets encountered in clinical practice.

Dose-response has been explored with multi-stent trabecular approaches. A 42-month study of one, two, or three iStent trabecular micro-bypass stents showed ≥ 20% IOP reduction without medication in 61% of one-stent eyes vs 91% of two- and three-stent eyes, suggesting that single-stent trabecular bypass provides initial efficacy but multi-stent approaches deliver more durable IOP control [2]. However, trabecular bypass remains constrained by the physiological floor imposed by episcleral venous pressure, with postoperative IOPs typically in the mid-to-upper teens [1].

The uveoscleral (unconventional) outflow pathway represents a physiologically distinct mechanism of aqueous humor drainage, estimated to account for approximately 35–50% of total aqueous outflow in healthy eyes. Unlike conventional trabecular outflow, which is strongly influenced by the pressure gradient across the trabecular meshwork and downstream episcleral venous pressure, uveoscleral outflow is relatively pressure-insensitive within the physiological IOP range. This pathway therefore provides an alternative route for aqueous drainage that is less dependent on conventional outflow resistance.

Importantly, conventional outflow is constrained by downstream resistance and episcleral venous pressure, which may impose a functional lower limit on IOP reduction, estimated at approximately 16 mmHg in a comprehensive Lancet review [4]. Uveoscleral drainage is not directly subject to this same episcleral venous pressure constraint, providing a distinct physiological rationale for therapeutic interventions targeting this pathway [3,4].

Bio-reinforced cyclodialysis is a novel conjunctiva-sparing approach to uveoscleral outflow enhancement. Because the procedure does not disturb the bulbar conjunctiva or Tenon’s capsule, conjunctival real estate remains preserved for subsequent bleb-forming filtration surgery (trabeculectomy, tube shunt, or subconjunctival microstent) should additional IOP-lowering be required. Allogeneic, acellular scleral bio- scaffolds reinforce the cyclodialysis cleft to create a direct communication between the anterior chamber and supraciliary space. We hypothesized that, like pharmacotherapy, the magnitude of uveoscleral outflow enhancement could be titrated by modulating the extent of the cyclodialysis cleft and the volume of bio-scaffold used to maintain it.

Using data from the prospective, multicenter CREST study (NCT05506423), we compared postoperative IOP, medication burden and safety between eyes receiving one versus two allogeneic scleral spacers during bio-reinforced cyclodialysis combined with cataract surgery. We hypothesized that eyes receiving two spacers would achieve greater 12-month IOP reduction than eyes receiving one. Unlike most angle- based surgical devices, which deliver a fixed intervention, spacer number is a parameter the surgeon can titrate intraoperatively [5]. Because spacer number was selected by the operating surgeon rather than assigned, the analysis was designed around methods that account for measured differences between the groups, and we report the extent to which any observed difference depends on those adjustments.

## Methods

### Study Design and Ethics

The CREST Study is a prospective, multicenter, observational study with a registered protocol, designed to function as a real-world study. Patients with open-angle glaucoma (OAG) who undergo bio- interventional uveoscleral enhancement surgery with the AlloFlo™ Uveo system (Iantrek, Inc., White Plains, NY, USA) are eligible to be followed through 24 months postoperatively. Inclusion criteria are deliberately broad (a diagnosis of OAG and an attempted bio-reinforced cyclodialysis procedure), and there are no exclusion criteria. The protocol specifies scheduled follow-up visits and assessments but imposes no preoperative medication washout and no postoperative medication or treatment policies.

These decisions, including the indication for surgery and the number of spacers placed, are left to the treating surgeon’s clinical judgment. Eyes are enrolled into Cohort 1 when the procedure is combined with phacoemulsification and into Cohort 2 when it is performed standalone. The study is conducted in accordance with the Declaration of Helsinki and was approved by the Advarra Institutional Review Board (IRB; protocol Pro00063287), which served as the central IRB, and by the Institutional Review Board at each participating site. All participants provided written informed consent.

## Study Population and Participant Flow

The present analysis includes 260 eyes (187 patients) that underwent bio-reinforced cyclodialysis combined with phacoemulsification cataract surgery (Cohort 1), received 1 or 2 spacers, and reached the month-12 study visit. Of these 260 eyes, 146 received one spacer and 114 received two. A total of 73 patients underwent bilateral cyclodialysis, contributing 146 eyes to the analysis set. Participant flow, including the 50 eyes that did not reach the month-12 visit, is shown in Fig. 1. Results are reported as of the most-recently reconciled data cutoff (31 January 2026).

**Fig. 1.** STROBE participant flow diagram showing derivation of the 260-eye analysis set from 577 records in the CREST database, with reasons for exclusion and the number of eyes contributing to each analysis. IOP intraocular pressure, STROBE Strengthening the Reporting of Observational Studies in Epidemiology

## Surgical Procedure

The bio-reinforced cyclodialysis procedure was performed ab-interno under gonioscopic visualization. A controlled cyclodialysis cleft was created in the nasal angle with the anterior chamber maintained by viscoelastic. Elongated 5-mm × 500-micron acellular scleral allograft bio-scaffolds (Fig. 2) were deployed into the supraciliary space, flush with the iris root, to maintain cleft patency and serve as a porous conduit for aqueous drainage into the uveoscleral pathway. Components do not protrude into the anterior chamber (AC-clear placement). Residual viscoelastic was evacuated with irrigation or irrigation/aspiration. Patients received a surgeon’s choice of postoperative topical antibiotic and corticosteroid regimen with taper.

**Fig. 2.** Bio-reinforced cyclodialysis: surgical concept and in situ imaging. A Bio-interventional cyclodialysis with the allograft spacer fully deployed to maintain a durable uveoscleral outflow conduit. B Processed micro-trephined scleral bio-scaffold made of porous, acellular, hydrophilic, collagenous sterile matrix. C Sleeved interventional deployment with a cyclodialysis cannula. D Gonioscopy view of the allogenic bio-spacer during intraocular implantation. E Post-implantation 90-day corneal OCT section showing the in situ allograft reinforcement. F Post-implantation 90-day longitudinal OCT imaging of the uveoscleral outflow-enhancing bio-scaffold maintaining a durable aqueous conduit and filtration reservoir. G External transluminating image demonstrating the visible allograft spacers at 90 days postoperatively. OCT optical coherence tomography

## Outcome Measures

Demographics, treatment history, and disease characteristics were recorded at enrollment. Post-surgical follow-up data were collected prospectively at months 1, 6 and 12, in addition to any unscheduled visits. Baseline IOP was the medicated IOP on the patient’s current regimen. Efficacy endpoints included change in IOP from baseline to month 12, the proportions of eyes achieving ≥ 20% and ≥ 30% IOP reduction, the proportions achieving month-12 IOP of ≤ 18 and ≤ 15 mmHg, and change in the number of IOP-lowering medications. Each IOP threshold was also evaluated as a composite that additionally required no increase in medication count from baseline and no secondary IOP-lowering procedure. In the composite, eyes that underwent a secondary procedure were classified as non-responders rather than censored. Baseline glaucoma severity was staged by visual field mean deviation (VF MD) using the Hodapp–Parrish–Anderson (HPA) convention as mild (MD > −6 dB), moderate (MD −6 to −12 dB), or severe (MD < −12 dB). The term “severe” is used throughout in preference to “advanced.”

Safety endpoints included clinically significant hypotony (IOP ≤ 5 mmHg with maculopathy), IOP elevation ≥ 10 mmHg from baseline at ≥ 30 days postoperatively, clinically significant hyphema, macular edema, corneal complications, and progression to secondary surgical intervention for IOP control. In addition, IOP elevation was summarized from all recorded IOP measurements within 12 months, scheduled and unscheduled and including the first postoperative month, as a rise of ≥ 10 mmHg above baseline or above the previous assessment.

## Handling of Secondary Procedures and Missing Data

IOP and medication values recorded after a secondary IOP-lowering procedure were censored, because such values reflect the subsequent intervention rather than the index procedure. Secondary IOP-lowering procedures were defined by procedure type irrespective of the recorded indication, and comprised laser trabeculoplasty, cyclophotocoagulation, goniotomy, canaloplasty, iridoplasty, synechiolysis, laser suture lysis, trabeculectomy, tube shunt and gel stent surgery, and bleb revision. Anterior chamber paracentesis, needling, neodymium-doped yttrium aluminum garnet (YAG) capsulotomy and intravitreal injections were not counted; sustained-release bimatoprost and travoprost implants were counted as medications rather than procedures. The primary analyses are as observed, without imputation, and the number of eyes contributing to every estimate is reported.

## Statistical Analysis

Baseline comparability was assessed using standardized mean difference (SMD), which expresses imbalance on a common scale that does not depend on sample size, with an absolute SMD above 0.10 taken to indicate meaningful imbalance. The primary endpoint was analyzed by five complementary approaches, specified before the adjusted results were examined: (1) unadjusted comparison of IOP change by Welch t-test; (2) analysis of covariance (ANCOVA) for month-12 IOP adjusted for baseline IOP; (3) ANCOVA additionally adjusted for baseline medication count, age, sex, glaucoma diagnosis and any prior glaucoma procedure; (4) 1:1 nearest-neighbor propensity-score matching without replacement, using a caliper of 0.2 standard deviations of the logit of the propensity score; and (5) overlap weighting with a doubly robust outcome model, which retains the full sample and weights each eye by its probability of receiving the alternative strategy.

The propensity model was a logistic regression of two-spacer treatment on baseline IOP, baseline medication count, age, sex, primary open-angle and pseudoexfoliative diagnoses, and any prior glaucoma procedure. Because spacer count proved to be strongly associated with enrolling site, two further analyses examined that structure directly: a linear mixed model with site as a random effect, and a fixed-effect model restricted to sites that used both strategies. A sensitivity analysis excluded the single highest- enrolling site. A Kaplan-Meier estimate of freedom from secondary incisional glaucoma surgery was performed with log-log CIs and log-rank comparison, as was a site-adjusted comparison of adverse-event rates.

Month-12 IOP, responder proportions and medication counts were summarized within each baseline severity stratum (mild, moderate, severe) by spacer number and are reported in Supplementary Table S3. Whether baseline severity predicted outcome, or modified the effect of spacer number, was examined in the main analysis by adding baseline visual field MD as a continuous covariate, and a spacer-by-MD interaction term, to the model for month-12 IOP adjusted for baseline IOP and enrolling site. Percentage IOP reduction was calculated for each eye relative to that eye’s own baseline and then averaged across eyes.

Because some eyes were bilateral treatment pairs, the between-eye correlation of IOP change was estimated and the mixed-model analyses were repeated with a patient random effect, both alone and together with site. Proportions were compared using Fisher exact-tests with Clopper-Pearson confidence intervals (CIs). Statistical significance was set at p < 0.05, and all tests were two-sided. No sample size or power calculation was performed, as the analysis used all eligible eyes in the registry. Analyses used R version 4.5.3 (R Foundation for Statistical Computing).

## Results

### Baseline Characteristics

Baseline characteristics are shown in Table 1. The two groups were similar in age (72.3 versus 72.4 years for one versus two spacers, respectively) and glaucoma diagnosis. 50.0% versus 56.1% were female and 50.0% versus 43.9% male. Two-spacer eyes were more often of Black race (36.0% versus 17.8%) and of Hispanic or Latino ethnicity (39.5% versus 21.2%); the ethnicity difference reflected enrolling site, as all eyes at the site contributing most two-spacer eyes were of Hispanic or Latino ethnicity. Baseline IOP was modestly higher in the two-spacer group (18.5 [5.7] versus 17.8 [4.2] mmHg; SMD 0.12), as was baseline medication count (1.5 [1.4] versus 1.3 [1.1]; SMD 0.15). Eyes receiving two spacers were less likely to have undergone a prior glaucoma procedure (11.4% versus 15.1%; SMD -0.11) and had worse visual field MD (-8.0 versus -6.2 dB; SMD -0.26). Among eyes with visual field data (N=87 one-spacer and N=59 two-spacer), glaucoma was mild in 62.1% versus 50.8%, moderate in 19.5% versus 22.0% and severe in 18.4% versus 27.1% (p = 0.350); severity at month 12 is shown in Supplementary Table S2.

**Table 1.** Baseline characteristics and standardized mean differences

| Characteristic | One spacer<br>(N = 146) | Two spacers<br>(N = 114) | SMD |
|---|---|---|---|
| Age, mean (SD), years | 72.3 (6.7) | 72.4 (7.9) | 0.00 |
| Female, n (%) | 73 (50.0) | 64 (56.1) | 0.12 |
| Male, n (%) | 73 (50.0) | 50 (43.9) | −0.12 |
| White, n (%) | 119 (81.5) | 73 (64.0) | −0.40 |
| Black or African American, n (%) | 26 (17.8) | 41 (36.0) | 0.42 |
| Asian, n (%) | 1 (0.7) | 0 (0.0) | −0.12 |
| Hispanic or Latino, n (%) | 31 (21.2) | 45 (39.5) | 0.40 |
| Primary open-angle glaucoma, n (%) | 145 (99.3) | 111 (97.4) | −0.15 |
| Pseudoexfoliative glaucoma, n (%) | 2 (1.4) | 0 (0.0) | −0.17 |
| Baseline IOP, mean (SD), mmHg | 17.8 (4.2) | 18.5 (5.7) | 0.12 |
| Baseline medications, mean (SD), n | 1.3 (1.1) | 1.5 (1.4) | 0.15 |
| Any prior glaucoma procedure, n (%) | 22 (15.1) | 13 (11.4) | −0.11 |
| Prior glaucoma procedures, mean (SD), n | 0.2 (0.5) | 0.2 (0.7) | 0.00 |
| Baseline visual field MD, mean (SD), dB□ | −6.2 (6.8) | −8.0 (7.6) | −0.26 |
| Baseline glaucoma severity<sup>a</sup> |  |  |  |
| Mild, n (%) | 54 (62.1) | 30 (50.8) | −0.23 |
| Moderate, n (%) | 17 (19.5) | 13 (22.0) | 0.06 |
| Severe, n (%) | 16 (18.4) | 16 (27.1) | 0.21 |
| Bilateral procedure, n (%) | 79 (54.1) | 67 (58.8) | 0.09 |
| Enrolling Site 9, n (%) | 11 (7.5) | 44 (38.6) | 0.79 |

IOP = intraocular pressure, MD = mean deviation, SD = standard deviation, SMD = standardized mean difference.
a. Baseline visual field data available for 87 one-spacer and 59 two-spacer eyes; percentages derived using these denominators. Severity characterized according to Hodapp-Parrish-Anderson (HPA) staging: mild, MD better than −6 dB; moderate, −6 to −12 dB; severe, worse than −12 dB.

The largest imbalance by a wide margin was enrolling site. One site contributed 44 of 114 two-spacer eyes (38.6%) but only 11 of 146 one-spacer eyes (7.5%), an SMD of 0.79. Three sites contributed 66 eyes that received one spacer exclusively. When enrolling site was included in the propensity model, 67 of 252 eyes (27%) fell outside the region of common support, having a fitted probability of treatment below 0.01 or above 0.99. Spacer count and site are therefore only partially separable in this dataset.

## IOP at Month 12

Month-12 IOP was available for 252 eyes (181 patients) after censoring (N=141 one-spacer, N=111 two- spacer). Of the 58 eyes without an analyzable month-12 IOP, 8 were censored after a secondary IOP- lowering procedure and 50 had no month-12 assessment: 14 had completed the study early, 8 relocated, 7 were lost to follow-up, 2 were non-adherent, 1 discontinued because of an adverse event and the remaining 18 were recorded as other or not stated. Baseline IOP did not differ significantly between eyes with and without month-12 data in either group (one spacer 17.8 versus 18.2 mmHg, p = 0.692; two spacers 18.4 versus 19.9 mmHg, p = 0.253). IOP fell from 17.8 to 14.9 mmHg with one spacer, a reduction of 2.9 mmHg (13.5%), and from 18.4 to 13.0 mmHg with two spacers, a reduction of 5.5 mmHg (23.6%).

A summary of efficacy outcomes is shown in Table 4. Effect estimates are shown in Table 2. The unadjusted between-group difference in IOP reduction was 2.5 mmHg (1.2, 3.8; p < 0.001). Adjustment for baseline IOP reduced this to 2.0 mmHg (1.2, 2.8), and adjustment for the full covariate set gave 2.0 mmHg (1.2, 2.8). Propensity-score matching produced 103 matched pairs with good balance on patient- level covariates (post-match SMD 0.06 for baseline IOP and -0.02 for medication count) and a difference of 2.4 mmHg (0.9, 3.8; p = 0.001). Overlap weighting with a doubly robust model, which retained all 252 eyes, gave 2.0 mmHg (1.2, 2.8; p < 0.001).

**Table 2.** Between-group difference in month-12 IOP reduction by analytical approach

| Analysis | Eyes, n | Difference, mmHg (95% CI)□ | Between-group p-value |
|---|---|---|---|
| Unadjusted (Welch t-test) | 252 | 2.5 (1.2, 3.8) | < 0.001 |
| ANCOVA, baseline IOP | 252 | 2.0 (1.2, 2.8) | < 0.001 |
| ANCOVA, full covariate set□ | 252 | 2.0 (1.2, 2.8) | < 0.001 |
| 1:1 propensity-score matching (103 pairs) | 206 | 2.4 (0.9, 3.8) | 0.001 |
| Overlap weighting, doubly robust | 252 | 2.0 (1.2, 2.8) | < 0.001 |
| Mixed model, enrolling site random effect□ | 252 | 0.9 (-0.1, 1.8) | 0.080 |
| Sites using both strategies only | 185 | 0.4 (-0.6, 1.5) | 0.424 |
| Excluding highest-enrolling site | 197 | 1.4 (0.5, 2.3) | 0.004 |
| Mixed model, patient random effect□ | 252 | 1.6 (0.8, 2.5) | < 0.001 |
| Mixed model, site and patient random effects□ | 252 | 0.7 (-0.2, 1.7) | 0.135 |

ANCOVA analysis of covariance, CI confidence interval, IOP intraocular pressure.
a. Positive values favor two spacers (greater IOP reduction).
b. Baseline IOP, baseline medication count, age, sex, glaucoma diagnosis, any prior glaucoma procedure.
c. Mixed models adjusted for baseline IOP and baseline medication count.

**Table 3.** Month-12 IOP change by enrolling site

| Site | One spacer, n | One spacer, change (mmHg) | Two spacers, n | Two spacers, change (mmHg) |
|---|---|---|---|---|
| Site 1 | 10 | −5.1 | 36 | −2.3 |
| Site 2 | 32 | −1.7 | 0 | – |
| Site 3 | 18 | −1.3 | 17 | −3.5 |
| Site 4 | 22 | −1.6 | 0 | – |
| Site 5 | 12 | −3.1 | 0 | – |
| Site 6 | 15 | −2.1 | 1 | −9.0 |
| Site 7 | 21 | −4.8 | 12 | −4.1 |
| Site 8 | 0 | – | 1 | +1.0 |
| Site 9 | 11 | −7.3 | 44 | −9.2 |

IOP, intraocular pressure.
Eyes with paired baseline and month-12 IOP after censoring at secondary procedures.

**Table 4.** Month-12 IOP and medication outcomes

| Outcome | One spacer<br>(N=141) | Two spacers<br>(N=111) | Between-group p-value□ |
|---|---|---|---|
| Month-12 IOP, mean (SD) mmHg | 14.9 (3.3) | 13.0 (3.3) | < 0.001 |
| Absolute IOP reduction, mean (95% CI) mmHg | 2.9 (2.2, 3.7) | 5.5 (4.3, 6.6) | < 0.001 |
| Relative IOP reduction, mean (95% CI) % | 13.5 (9.8, 17.1) | 23.6 (18.5, 28.7) | 0.002 |
| Reduction of 20% or more, n (%) | 54 (38.3) | 65 (58.6) | 0.002 / 0.476 |
| With no medication increase or secondary procedure□ | 47/146 (32.2) | 60/114 (52.6) | < 0.001 / 0.280 |
| Reduction of 30% or more, n (%) | 34 (24.1) | 53 (47.7) | < 0.001 / 0.247 |
| With no medication increase or secondary procedure□ | 31/146 (21.2) | 49/114 (43.0) | < 0.001 / 0.243 |
| IOP 18 mmHg or below, n (%) | 120 (85.1) | 104 (93.7) | 0.042 / 0.423 |
| With no medication increase or secondary procedure□ | 112/146 (76.7) | 99/114 (86.8) | 0.039 / 0.288 |
| IOP 15 mmHg or below, n (%) | 84 (59.6) | 89 (80.2) | < 0.001 / 0.089 |
| With no medication increase or secondary procedure□ | 79/146 (54.1) | 85/114 (74.6) | < 0.001 / 0.044 |
| IOP 12 mmHg or below, n (%) | 32 (22.7) | 50 (45.0) | < 0.001 / 0.005 |
| With no medication increase or secondary procedure□ | 30/146 (20.5) | 49/114 (43.0) | < 0.001 / < 0.001 |
| Medication change, mean (95% CI) n | –0.7 (–0.9, –0.5) | –0.6 (–0.8, –0.4) | 0.643 |
| Medication-free at month 12, % | 58.2 | 51.4 | 0.309 |

CI confidence interval, IOP intraocular pressure.
IOP and medication values recorded after a secondary IOP-lowering procedure were censored.
a. For responder rows, unadjusted Fisher exact p-value / p-value from mixed-effects logistic regression with enrolling site as a random effect.
b. Composite: criterion met with no increase in medication count from baseline and no secondary IOP-lowering procedure; eyes undergoing a secondary procedure are counted as non-responders.

Matching on patient-level covariates did not balance enrolling site, which remained imbalanced after matching (SMD 0.71). When site was accounted for, the estimate attenuated substantially: 0.9 mmHg in a mixed model with site as a random effect (-0.1, 1.8; p = 0.080), and 0.4 mmHg when the analysis was restricted to the five sites that used both strategies (-0.6, 1.5; p = 0.424). Excluding the highest-enrolling site gave 1.4 mmHg (0.5, 2.3; p = 0.004). Site-level results are shown in Table 3: mean IOP reduction ranged from 1.3 to 7.3 mmHg among one-spacer eyes and from 2.3 to 9.2 mmHg among two-spacer eyes, and the site contributing most two-spacer eyes had both the highest baseline IOP (21.8 mmHg) and the largest reduction (9.2 mmHg).

IOP change was strongly correlated between fellow eyes (r = 0.76; 0.65, 0.85; N=71 patients). Accounting for this did not alter the findings: with a patient random effect and adjustment for baseline IOP and medications the difference was 1.6 mmHg (0.8, 2.5; p < 0.001), and with both site and patient random effects it was 0.7 mmHg (-0.2, 1.7; p = 0.135) (Table 2).

## Responders and Medications at Month 12

In unadjusted analyses, a greater proportion of two-spacer eyes achieved at least a 20% IOP reduction (58.6% [48.8, 67.8] versus 38.3% [30.2, 46.9]; p = 0.002) and at least a 30% reduction (47.7% [38.2, 57.4] versus 24.1% [17.3, 32.0]; p < 0.001). Month-12 IOP of 15 mmHg or below was achieved by 80.2% versus 59.6% (p < 0.001) and 12 mmHg or below by 45.0% versus 22.7% (p < 0.001). When each criterion additionally required no increase in medications and no secondary IOP-lowering procedure, rates were lower in both groups but the pattern was unchanged; for example, 52.6% versus 32.2% achieved at least a 20% reduction and 74.6% versus 54.1% reached 15 mmHg or below (Table 4). After adjustment for enrolling site, between-group differences in responder rates were attenuated: they were no longer statistically significant for the 20% (OR 1.29; 0.64, 2.59) and 30% reduction criteria or for IOP of 18 mmHg or below, were borderline for 15 mmHg or below (OR 2.04; 0.90, 4.65), and remained significant for 12 mmHg or below (OR 2.79; 1.36, 5.74). IOP over time is shown in Fig. 3.

**Fig. 3.** IOP over time through month 12, by spacer count. Data are presented as mean (95% CI). Denominators at baseline, month 1, month 6 and month 12 were 146, 146, 128 and 141 for one spacer and 114, 113, 90 and 111 for two spacers. IOP recorded after a secondary IOP-lowering procedure was censored. CI confidence interval, IOP intraocular pressure

Paired baseline and month-12 medication counts were available for 252 eyes (141 one-spacer, 111 two- spacer). Medication count decreased by 0.7 (0.5, 0.9; p < 0.001) from 1.3 ± 1.1 at baseline to 0.6 ± 0.9 with one spacer, and by 0.6 (0.4, 0.8; p < 0.001) from 1.5 ± 1.4 to 0.9 ± 1.2 with two spacers. The between-group difference was not significant (p = 0.643). The proportion of medication-free eyes rose from 24.8% to 58.2% with one spacer and from 23.4% to 51.4% with two.

## Outcomes by Baseline Glaucoma Severity

Baseline visual-field data were available for 146 of 260 eyes, of which 139 (83 one-spacer, 56 two- spacer) had an analyzable month-12 IOP after censoring. In a model for month-12 IOP adjusted for baseline IOP and enrolling site, baseline visual field MD was not predictive of month-12 IOP (0.06 mmHg per dB; −0.02, 0.14; p = 0.154), and there was no interaction between spacer number and baseline MD (p = 0.366). Outcomes within the mild, moderate, and severe strata are shown in Supplementary Table S3. The between-group difference in IOP reduction was close to zero in all three (0.2, 0.0 and −0.4 mmHg). Visual field availability was itself strongly related to site: at the site contributing most two- spacer eyes, only 1 of 44 had a baseline visual field, and among the remaining centers one and two spacers did not differ (0.1 mmHg; −1.5, 1.8; p = 0.880). This analysis is therefore largely a within-center comparison rather than an independent test of effect modification.

## Safety Outcomes

Safety and adverse events are summarized in Table 5. A secondary IOP-lowering procedure within the 12-month window was performed in 5 of 146 one-spacer eyes (3.4%; 1.1, 7.8) and 4 of 114 two-spacer eyes (3.5%; 1.0, 8.7; p = 1.000); three one-spacer and three two-spacer eyes underwent laser alone, two one-spacer eyes underwent incisional surgery (gel stent and tube shunt), and one two-spacer eye underwent cyclophotocoagulation combined with goniotomy. Kaplan–Meier freedom from secondary incisional glaucoma surgery at 12 months was 98.6% (94.6, 99.7) with one spacer and 99.1% (93.9, 99.9) with two (log-rank p = 0.669). Any ocular adverse event was recorded in 22 one-spacer eyes (15.1%) and 10 two-spacer eyes (8.8%; p = 0.133). There were no cases of clinically significant hypotony in either group and no cases of spacer dislodgement, migration or extrusion.

**Table 5.**
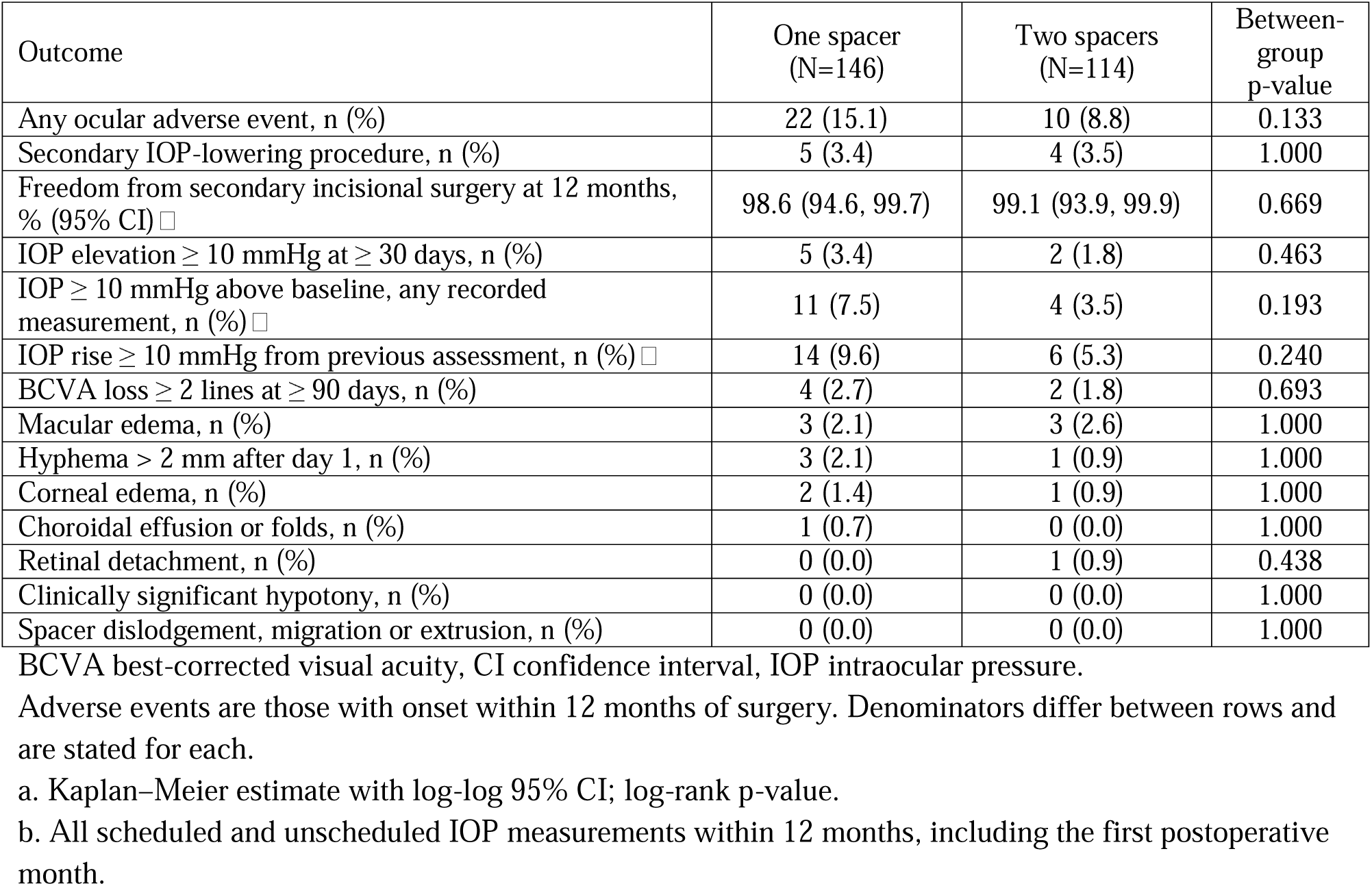
Summary of Safety

| Outcome | One spacer<br>(N=146) | Two spacers<br>(N=114) | Between-group p-value |
|---|---|---|---|
| Any ocular adverse event, n (%) | 22 (15.1) | 10 (8.8) | 0.133 |
| Secondary IOP-lowering procedure, n (%) | 5 (3.4) | 4 (3.5) | 1.000 |
| Freedom from secondary incisional surgery at 12 months, % (95% CI)□ | 98.6 (94.6, 99.7) | 99.1 (93.9, 99.9) | 0.669 |
| IOP elevation ≥ 10 mmHg at ≥ 30 days, n (%) | 5 (3.4) | 2 (1.8) | 0.463 |
| IOP ≥ 10 mmHg above baseline, any recorded measurement, n (%)□ | 11 (7.5) | 4 (3.5) | 0.193 |
| IOP rise ≥ 10 mmHg from previous assessment, n (%)□ | 14 (9.6) | 6 (5.3) | 0.240 |
| BCVA loss ≥ 2 lines at ≥ 90 days, n (%) | 4 (2.7) | 2 (1.8) | 0.693 |
| Macular edema, n (%) | 3 (2.1) | 3 (2.6) | 1.000 |
| Hyphema > 2 mm after day 1, n (%) | 3 (2.1) | 1 (0.9) | 1.000 |
| Corneal edema, n (%) | 2 (1.4) | 1 (0.9) | 1.000 |
| Choroidal effusion or folds, n (%) | 1 (0.7) | 0 (0.0) | 1.000 |
| Retinal detachment, n (%) | 0 (0.0) | 1 (0.9) | 0.438 |
| Clinically significant hypotony, n (%) | 0 (0.0) | 0 (0.0) | 1.000 |
| Spacer dislodgement, migration or extrusion, n (%) | 0 (0.0) | 0 (0.0) | 1.000 |

BCVA best-corrected visual acuity, CI confidence interval, IOP intraocular pressure.
Adverse events are those with onset within 12 months of surgery. Denominators differ between rows and are stated for each.
a. Kaplan–Meier estimate with log-log 95% CI; log-rank p-value.
b. All scheduled and unscheduled IOP measurements within 12 months, including the first postoperative month.

No statistically significant difference in observed adverse-event rates was detected; however, the study was not powered to establish safety equivalence. With 146 and 114 eyes per group and no observed events in a category, the rule of three permits exclusion only of event rates above approximately 2.1% and 2.6%, respectively. The proportion of eyes with a recorded adverse event varied between sites, from 0% to 41.7% among sites contributing more than one eye (Supplementary Table S1). After accounting for site, the odds of any ocular adverse event did not differ between groups (OR 0.72; 0.27, 1.94; p = 0.523), and when the site contributing most two-spacer eyes was excluded rates were comparable between groups (16.3% and 14.3%; p = 0.840). The between-group safety comparison should therefore be regarded as descriptive. For reference, across all 310 enrolled eyes, including the 50 that did not reach the month-12 visit, any ocular adverse event was recorded in 12.9% of one-spacer and 7.9% of two-spacer eyes.

Because study visits began at month 1, transient early IOP elevation could be ascertained only from unscheduled visits: 130 of 260 eyes (50.0%) had at least one IOP measurement in the first 30 days and 24 (9.2%) in the first 14 days. Considering all recorded measurements within 12 months, an IOP ≥ 10 mmHg above baseline was recorded in 11 one-spacer eyes (7.5%) and 4 two-spacer eyes (3.5%; p = 0.193), including 8 and 2 eyes, respectively, in the first 30 days. A rise of ≥ 10 mmHg from the previous assessment was recorded in 14 (9.6%) and 6 (5.3%) eyes (p = 0.240). These figures are likely to underestimate the incidence of early transient IOP spikes.

No best-corrected visual acuity (BCVA) loss of 2 or more lines recorded at 90 days or later within the 12- month window (n=6) was adjudicated as related to the cyclodialysis procedure or to the bio-scaffold; one case (in a single-spacer eye) was attributed to the intraocular lens (IOL) and recovered after YAG capsulotomy, two were attributed to comorbid retinal disease (branch retinal vein occlusion with macular edema; diabetic macular edema) and three had no comorbidity recorded and resolved without intervention. Three further BCVA-loss events occurred after month 12 and fall outside the reporting window; none was adjudicated as related to the procedure or the bio-scaffold, and one had no completed adjudication.

Macular edema cases (n=6) were predominantly cystoid macular edema of presumed pseudophakic origin; none were adjudicated as related to the bio-scaffold. Five resolved within 12 weeks of onset; 1 remained unresolved at last follow-up.

One unrelated retinal detachment resolved with standard of care. One choroidal folds event, adjudicated as “probably” related to the bio-scaffold, presented at month 1 and resolved without sequelae approximately 7.5 months later after viscoelastic reformation of the anterior chamber and argon laser treatment.

Unplanned ocular procedures recorded within the 12-month window comprised neodymium-doped yttrium aluminum garnet (YAG) capsulotomy (n=20), intravitreal injection (n=6), other retinal surgery (n=2) and removal of a cortical remnant (n=1); none were adjudicated as related to the bio-scaffold, and all resolved without sequelae. All postoperative hyphemas resolved with conservative management.

Transient numeric hypotony (IOP < 6 mmHg without maculopathy) cases all resolved spontaneously.

## Discussion

In this multicenter real-world cohort of 260 eyes undergoing combined phacoemulsification and bio- reinforced cyclodialysis, implantation of two scleral allograft spacers was associated with greater intraocular pressure (IOP) reduction at 12 months than implantation of one. The unadjusted difference in IOP reduction was 2.5 mmHg, and an approximately 2-mmHg association persisted after adjustment for baseline IOP and other patient-level covariates, propensity-score matching, and doubly robust overlap weighting. The consistency of these estimates suggests that the observed gradient was not readily explained by measured differences in patient characteristics alone. However, spacer number was strongly associated with enrolling center. Once center was accounted for, the estimated difference attenuated to 0.9 mmHg in a mixed-effects model and to 0.4 mmHg when the analysis was restricted to centers using both strategies.

These findings suggest a numerical trend toward greater IOP reduction with increasing spacer number, although the independent contribution of spacer number could not be reliably distinguished from intercenter variability. Accordingly, the absence of a statistically significant site-adjusted effect should not be interpreted as evidence of biological equivalence between single- and dual-spacer implantation. The confidence interval surrounding the adjusted estimate remained compatible with a potentially clinically meaningful incremental benefit, while sensitivity analysis excluding the highest-enrolling center demonstrated a 1.4-mmHg difference. Collectively, these observations support the plausibility of a spacer-number–dependent effect but remain insufficient to establish an independent dose–response relationship. Adjustment for intereye correlation in patients contributing both eyes did not materially alter these findings.

## Titratability as a Concept in Minimally Invasive Glaucoma Surgery

The ability to titrate the magnitude of a glaucoma intervention has important clinical implications. Medical therapy is routinely escalated according to baseline disease severity, target IOP, achieved pressure, risk of progression, medication tolerance, and treatment response. In contrast, many surgical interventions are delivered as a relatively fixed treatment. A surgical platform in which treatment intensity can be modified intraoperatively could extend the principles of individualized glaucoma management to surgery.

Trabecular micro-bypass provides the most direct clinical precedent. In a prospective randomized study of 119 eyes assigned to implantation of one, two, or three first-generation iStents as a standalone procedure, mean unmedicated IOP at 18 months was 15.9, 14.1, and 12.2 mmHg, respectively, each additional stent producing a statistically significant incremental reduction [6]. The gradient persisted at approximately 42 months, when post-washout IOP was 17.4, 15.8, and 14.2 mmHg and a ≥20% unmedicated reduction was maintained in 61% of one-stent eyes compared with 91% of two- and three-stent eyes, with medication use increasing over time predominantly in the single-stent group [2].

These observations are particularly relevant because titration may manifest in more than one dimension. Increasing surgical intensity may increase mean pressure reduction, increase the probability of reaching a lower target IOP, reduce medication dependence, improve durability, or some combination of these outcomes. In the present CREST cohort, two-spacer eyes similarly demonstrated higher unadjusted rates of ≥20% and ≥30% IOP reduction and substantially higher proportions reaching IOP thresholds of ≤15 and ≤12 mmHg. These responder differences, however, were attenuated after adjustment for site, although the proportion reaching 12 mmHg or below remained higher with two spacers; they should therefore be considered hypothesis-generating rather than evidence of an independent spacer-dose effect.

Non-randomized experience points the same way but is weaker. In 53 eyes undergoing phacoemulsification with either two or three iStents, 12-month IOP reduction was similar between groups while medication burden was lower with three stents (46% versus 72% medication-free), illustrating that incremental treatment intensity may be expressed through medication use even when mean IOP separation is small [7]. A cross-study comparison likewise reported progressively greater IOP reduction with phacoemulsification alone, one iStent, and two iStents, although it aggregates different studies and populations and is not a randomized dose comparison [8].

## Biological Plausibility: Number Versus Extent of Outflow Enhancement

Experimental work provides a mechanistic basis and suggests that the more fundamental variable may be the extent of the outflow pathway modified rather than device number itself. In perfused human anterior segments, a second iStent inject increased outflow facility beyond that achieved with one [9]. An 8-mm Schlemm’s canal scaffold, however, increased outflow facility by 74% compared with 34% after two focal trabecular micro-bypasses, a difference attributed in part to the greater length of canal dilation [10], and in a paired-eye human anterior segment perfusion model the same scaffold reduced outflow resistance most in the eyes with the highest baseline resistance [11].

This distinction may be especially relevant to bio-reinforced cyclodialysis. Trabecular micro-bypass creates discrete points of access to Schlemm’s canal, whereas cyclodialysis establishes communication between the anterior chamber and supraciliary space and reinforces that newly created pathway with an allogeneic scleral scaffold. The physiological pathways are different, but the broader principle may be analogous: modifying a greater effective extent or capacity of an aqueous outflow pathway could alter hydraulic conductance and thereby influence IOP.

For cyclodialysis, therefore, the relevant surgical “dose” may be more complex than spacer count alone. It may encompass circumferential cleft length, scaffold volume, the number and spatial distribution of reinforced segments, the effective surface area of communication with the supraciliary compartment, tissue characteristics, postoperative healing, and persistence of cleft architecture. Two spacers should consequently not be interpreted as simply twice the treatment delivered by one spacer. Spacer number may instead serve as a surrogate for a broader anatomical construct: the extent of reinforced cyclodialysis.

This distinction also helps reconcile the present findings with the iStent literature. Device count is a relatively discrete exposure in trabecular micro-bypass. In cyclodialysis, creation of the cleft precedes and accompanies scaffold implantation, and spacer number may correlate with surgeon-specific differences in cleft extent and technique. This makes a spacer-number effect inherently more difficult to isolate in a surgeon-selected observational registry and further supports prospective measurement of cleft anatomy in future studies.

## Why Enrolling Site Is Central to Interpretation

The dominant methodological feature of this study was the strong relationship between spacer strategy and enrolling center. One center contributed 38.6% of all two-spacer eyes but only 7.5% of one-spacer eyes, while three centers used a single spacer exclusively. When enrolling site was incorporated into the propensity model, 27% of eyes fell outside the region of common support. For those eyes, the dataset contains little or no credible within-site counterfactual comparison.

This represents more than ordinary residual confounding. When treatment selection is highly clustered within centers, patient-level propensity methods can produce excellent balance in measured clinical characteristics while leaving practice-level determinants of outcome unresolved. The persistence of an approximately 2-mmHg association after multiple patient-level adjustments therefore demonstrates robustness to measured case mix, but not to center-level variation.

Potential center effects include patient selection, target IOP, surgeon experience, the location and extent of the cyclodialysis cleft, spacer positioning, intraoperative manipulation, viscoelastic management, perioperative medications, corticosteroid regimens, postoperative management, thresholds for restarting medications or performing additional intervention, and measurement practices. Of particular relevance, the center contributing the greatest number of two-spacer eyes also had the highest baseline IOP and the largest mean IOP reduction.

Accordingly, attenuation after adjustment for site should not be interpreted as demonstrating that spacer number is biologically inert. It demonstrates that this dataset cannot identify its independent effect with sufficient precision. Conversely, the sizeable unadjusted and patient-adjusted differences cannot establish causality. Taken together, the two observations define the principal uncertainty of the study and provide a compelling rationale for randomized evaluation.

The severity analysis illustrates the same constraint from a different direction. Because the site contributing most two-spacer eyes recorded few baseline visual fields, the eyes available for stratification come largely from the remaining centers, and within that subset the unadjusted spacer difference was null.

The absence of a severity-by-spacer interaction therefore carries limited weight as evidence against effect modification in severe disease; it is better read as a further demonstration that the apparent spacer gradient in this dataset is inseparable from center.

## Relationship to Concomitant Cataract Surgery

All eyes underwent concomitant phacoemulsification, and no phacoemulsification-only comparator was included. The study therefore cannot determine the absolute IOP-lowering contribution of bio-reinforced cyclodialysis relative to cataract extraction alone. However, phacoemulsification was common to both spacer groups, so the principal study question remains the relative outcome of one- versus two-spacer strategies within the same combined-surgery setting [12].

The effect of cataract surgery remains relevant because IOP reduction following phacoemulsification is itself dependent on baseline pressure, reinforcing the importance of the multiple baseline-IOP adjustments performed in this analysis [13]. Regression to the mean is a related consideration: the site contributing most two-spacer eyes also had the highest mean baseline IOP, and eyes with higher measured baseline pressure tend to show larger subsequent reductions independent of treatment. Adjustment for baseline IOP mitigates, but cannot fully remove, this effect when baseline IOP and site are themselves associated.

The broader minimally invasive glaucoma surgery (MIGS) literature also illustrates the importance of distinguishing treated mean IOP from medication-independent efficacy. In the pivotal randomized iStent inject trial, phacoemulsification plus two iStent inject devices produced significantly greater unmedicated diurnal IOP reduction at 24 months than phacoemulsification alone (7.0 versus 5.4 mmHg; P<0.001), with ≥20% reductions in 75.8% versus 61.9% of eyes [14].

Conversely, longer-term Schlemm’s canal MIGS data demonstrate that meaningful medication-sparing effects may persist even when treated mean IOPs become similar. At 3 years in the randomized HORIZON trial, mean treated IOP was 16.7 mmHg with Hydrus plus cataract surgery versus 17.0 mmHg with cataract surgery alone (P=0.85), while mean medication use was 0.4 versus 0.8 and 73% versus 48% of eyes were medication-free [15]. Network meta-analysis of Schlemm’s canal and trabecular bypass approaches combined with phacoemulsification supports the same distinction between treated pressure and medication burden [16]. This distinction reinforces the importance of evaluating mean IOP, target attainment, medication burden, and durability as complementary, not interchangeable, measures of the clinical effect of titratable glaucoma surgery.

## Clinical Implications

The present findings define a testable clinical hypothesis rather than a definitive dose-response relationship. A prospective trial randomizing eyes to one versus two spacers within participating surgeons and centers would directly uncouple treatment intensity from surgeon preference, center practice, and case selection. Stratification by baseline IOP and glaucoma severity would further reduce important sources of heterogeneity.

Future studies should also move beyond spacer count alone. Prospective measurement of cyclodialysis cleft length and circumferential arc, spacer location and separation, scaffold volume, and postoperative anatomic persistence using anterior-segment imaging could identify the anatomical parameters most closely associated with physiological response. Such an approach would test whether the relevant variable is the number of scaffolds, the extent of reinforced uveoscleral communication, or both.

If a reproducible relationship between anatomic treatment extent and clinical response is demonstrated, bio-reinforced cyclodialysis could provide a framework for patient-specific titration of uveoscleral surgery, conceptually analogous to the graded effects observed with increasing trabecular micro-bypass treatment. The randomized iStent data demonstrate that surgical titratability is achievable within MIGS; the present CREST data extend that question to a physiologically distinct outflow pathway.

## Safety

Observed safety outcomes were favorable in both groups. Secondary IOP-lowering procedures occurred in 3.4% and 3.5% of eyes, respectively, and no cases of clinically significant hypotony or spacer dislodgement, migration, or extrusion were recorded, consistent with a previously reported series of more than 240 bio-reinforced cyclodialysis cases [17]. These observations should nevertheless be interpreted conservatively. The study was not designed or powered to establish safety equivalence, and uncommon complications cannot be excluded with the available sample size. Recorded adverse-event rates also varied considerably between sites, consistent with differences in reporting practice in a real-world setting; after accounting for site, they did not differ between groups. In addition, because the protocol had no day- 1 or week-1 visits, transient early IOP spikes, which after angle and supraciliary surgery occur predominantly in the first postoperative days to weeks, were incompletely captured. Accordingly, these findings should be viewed as descriptive rather than evidence that one- and two-allograft strategies have equivalent safety.

## Limitations and Strengths

Several limitations determine the boundaries of inference. First, spacer number was selected by the operating surgeon and was strongly associated with enrolling center, resulting in limited treatment overlap and preventing reliable separation of spacer and center effects. Second, cleft extent was not measured independently of spacer number, so scaffold count cannot be distinguished from other dimensions of surgical intensity. Third, all eyes underwent concomitant phacoemulsification without a cataract-surgery- only control, precluding estimation of the absolute treatment contribution of bio-reinforced cyclodialysis. Fourth, the analysis population was restricted to eyes reaching the month-12 visit, and a further eight measurements were censored after secondary IOP-lowering procedures; composite endpoints classified eyes requiring a secondary procedure as failures. Fifth, baseline glaucoma severity and 12-month IOP measurements were available for only N=139 eyes, yielding imprecise stratified estimates. Additionally visual-field availability was strongly related to site, so the severity-stratified analysis is largely a within- site comparison rather than an independent test of effect modification. Sixth, the study was not prospectively powered for the one-versus-two-spacer comparison or for safety equivalence. Seventh, the visit schedule did not include day-1 or week-1 visits, so early transient IOP elevations were captured only at unscheduled visits, and adverse-event recording varied between sites. Eighth, as in routine practice there was no medication washout, so IOP was measured on medication and medication changes were at the surgeon’s discretion. Finally, follow-up is limited to 12 months, and these are short-term results.

These limitations are balanced by several strengths. The analysis derives from a relatively large prospective, multicenter real-world cohort; uses a reconciled patient-level dataset with explicitly defined denominators; evaluates the primary association using multiple complementary statistical approaches; demonstrates highly concordant estimates across several methods of patient-level adjustment; and, importantly, interrogates rather than obscures the strong relationship between treatment strategy and center. The analysis therefore characterizes both the magnitude of the observed clinical signal and the limits of causal inference inherent in surgeon-selected real-world treatment.

## Conclusions

In this prospective multicenter real-world cohort, two-spacer bio-reinforced cyclodialysis was associated with numerically greater 12-month IOP reduction than one-spacer treatment in unadjusted and multiple patient-level adjusted analyses. An approximately 2-mmHg association persisted across covariate adjustment, propensity-score matching, and doubly robust overlap weighting, but was substantially attenuated when enrolling center was accounted for because spacer strategy was strongly clustered by site.

These findings identify a potential incremental treatment effect but do not establish that a second spacer independently causes additional IOP lowering. The hypothesis is supported by biological plausibility and by precedent from trabecular MIGS: randomized clinical data demonstrate graded IOP effects with increasing numbers of trabecular micro-bypass stents, while experimental studies indicate that outflow enhancement can scale with the extent of the pathway modified.

A prospective randomized comparison of one versus two spacers, incorporating standardized surgical technique and quantitative characterization of cyclodialysis cleft anatomy, is warranted to determine whether bio-reinforced cyclodialysis can provide a reproducible titratable uveoscleral surgical effect.

## Data Availability

The datasets generated during and/or analyzed during the current study are available from the corresponding author on reasonable request.

## Acknowledgments

We thank the participants of the study.

## Medical Writing and Editorial Assistance

No medical writing or editorial assistance was received in the preparation of this article.

## Authorship

All named authors meet the International Committee of Medical Journal Editors (ICMJE) criteria for authorship of this article, take responsibility for the integrity of the work as a whole, and have given their approval for this version to be published.

## Author Contributions

Conceptualization: Tsontcho Ianchulev. Methodology and formal analysis: Tsontcho Ianchulev. Investigation and data acquisition: Craig Chaya, Nathan M Radcliffe, Mary Qiu, Sandra Sieminski, Shamil Patel, Gabriel Ferreira, Manjool Shah and Jason Bacharach. Writing, original draft preparation: Tsontcho Ianchulev. Writing, review and editing: all authors. Supervision: Tsontcho Ianchulev. All authors read and approved the final manuscript.

## Ethical Approval

This study was performed in accordance with the ethical standards of the 1964 Declaration of Helsinki and its later amendments. The CREST study protocol was approved by the Advarra Institutional Review Board (IRB; protocol Pro00063287; OHRP/FDA registration IRB00000971), which served as the central IRB of record (approval number MOD02608603). All participants provided written informed consent before enrollment.

