## Supplemental Tables S1, S2, S3 for "Clinical outcomes of bio-interventional cyclodialysis with one or multiple allogeneic reinforcements: a multicenter real-world evidence study"

**Cover Page**

**Electronic Supplementary Material**

**Supplementary Table S1. Eyes with any recorded ocular adverse event within 12 months, by enrolling site**

| Site | One Spacer, n/N (%) | Two Spacers, n/N (%) | All Eyes, n/N (%) |
| --- | --- | --- | --- |
| Site 1 | 0/10 (0.0) | 5/39 (12.8) | 5/49 (10.2) |
| Site 2 | 4/32 (12.5) | – | 4/32 (12.5) |
| Site 3 | 8/21 (38.1) | 4/17 (23.5) | 12/38 (31.6) |
| Site 4 | 2/22 (9.1) | – | 2/22 (9.1) |
| Site 5 | 5/12 (41.7) | – | 5/12 (41.7) |
| Site 6 | 2/17 (11.8) | 0/1 (0.0) | 2/18 (11.1) |
| Site 7 | 1/21 (4.8) | 0/12 (0.0) | 1/33 (3.0) |
| Site 8 | – | 1/1 (100.0) | 1/1 (100.0) |
| Site 9 | 0/11 (0.0) | 0/44 (0.0) | 0/55 (0.0) |
| All sites | 22/146 (15.1) | 10/114 (8.8) | 32/260 (12.3) |
| All sites, excluding 9 | 22/135 (16.3) | 10/70 (14.3) | 32/205 (15.6) |

CI = confidence interval; OR = odds ratio.
Notes: Site numbering corresponds to Table 3. Between-group comparison accounting for site: mixed-effects logistic regression with site as a random effect, OR 0.72 (95% CI: 0.27, 1.94; p = 0.523); Mantel–Haenszel exact OR across the sites using both strategies 0.93 (95% CI: 0.29, 3.00).

**Supplementary Table S2. Glaucoma severity by visual field mean deviation at baseline and month 12**

|  | One spacer, n (%) | Two spacers, n (%) |
| --- | --- | --- |
| Baseline, eyes with visual field | N = 87 | N = 59 |
| Mild (MD better than −6 dB) | 54 (62.1) | 30 (50.8) |
| Moderate (−6 to −12 dB) | 17 (19.5) | 13 (22.0) |
| Severe (worse than −12 dB) | 16 (18.4) | 16 (27.1) |
| Mean MD, dB | -6.2 | -8.0 |
| Month 12, eyes with visual field | N = 125 | N = 52 |
| Mild (MD better than −6 dB) | 93 (74.4) | 34 (65.4) |
| Moderate (−6 to −12 dB) | 16 (12.8) | 5 (9.6) |
| Severe (worse than −12 dB) | 16 (12.8) | 13 (25.0) |
| Mean MD, dB | -4.7 | -7.1 |
| Eyes with paired baseline and month-12 fields | N = 80 | N = 44 |
| Mean MD change, dB | +1.0 | +0.6 |

MD = mean deviation.
Distribution of severity did not differ between groups at baseline (p = 0.350) or month 12 (p = 0.131; chi-square). Month-12 values are from different, partly overlapping sets of eyes and are not a measure of within-eye change.

**Supplementary Table S3. Month-12 IOP and medication outcomes by baseline glaucoma severity and spacer number**

| Severity  Outcome | One spacer | Two spacers |
| --- | --- | --- |
| Mild (MD better than −6 dB) | n = 52 | n = 29 |
| Baseline IOP, mean (SD) mmHg | 17.9 (4.3) | 15.4 (4.6) |
| Month-12 IOP, mean (SD) mmHg | 15.9 (3.5) | 13.2 (4.0) |
| Absolute IOP reduction, mean (95% CI) mmHg | 2.1 (0.7, 3.5) | 2.3 (0.4, 4.1) |
| Between-group difference, mean (95% CI) mmHg^a^ | 0.2 (−2.1, 2.5); p = 0.862 | |
| Relative IOP reduction, mean (95% CI) % | 7.8 (0.8, 14.9) | 10.4 (−1.1, 21.9) |
| Reduction of 20% or more, n (%) | 16 (30.8) | 12 (41.4) |
| Reduction of 30% or more, n (%) | 10 (19.2) | 8 (27.6) |
| IOP 15 mmHg or below, n (%) | 24 (46.2) | 20 (69.0) |
| IOP 12 mmHg or below, n (%) | 9 (17.3) | 12 (41.4) |
| Baseline medications, mean (SD) n | 1.2 (1.0) | 1.3 (1.2) |
| Month-12 medications, mean (SD) n | 0.5 (0.8) | 0.9 (1.4) |
| Medication change, mean (95% CI) n | −0.6 (−0.8, −0.4) | −0.3 (−0.7, 0.0) |
| Moderate (−6 to −12 dB) | n = 16 | n = 13 |
| Baseline IOP, mean (SD) mmHg | 16.8 (4.8) | 16.9 (6.7) |
| Month-12 IOP, mean (SD) mmHg | 13.0 (3.3) | 13.1 (1.9) |
| Absolute IOP reduction, mean (95% CI) mmHg | 3.8 (1.5, 6.1) | 3.8 (0.0, 7.7) |
| Between-group difference, mean (95% CI) mmHg^a^ | 0.0 (−4.3, 4.4); p = 0.987 | |
| Relative IOP reduction, mean (95% CI) % | 18.5 (5.7, 31.3) | 15.1 (0.1, 30.1) |
| Reduction of 20% or more, n (%) | 10 (62.5) | 4 (30.8) |
| Reduction of 30% or more, n (%) | 6 (37.5) | 4 (30.8) |
| IOP 15 mmHg or below, n (%) | 14 (87.5) | 12 (92.3) |
| IOP 12 mmHg or below, n (%) | 6 (37.5) | 3 (23.1) |
| Baseline medications, mean (SD) n | 1.4 (1.4) | 1.2 (1.0) |
| Month-12 medications, mean (SD) n | 0.9 (1.0) | 1.3 (1.0) |
| Medication change, mean (95% CI) n | −0.5 (−1.3, 0.3) | 0.2 (−0.4, 0.7) |
| Severe (MD worse than −12 dB) | n = 15 | n = 14 |
| Baseline IOP, mean (SD) mmHg | 17.3 (4.2) | 14.9 (4.3) |
| Month-12 IOP, mean (SD) mmHg | 14.7 (3.5) | 12.6 (4.3) |
| Absolute IOP reduction, mean (95% CI) mmHg | 2.7 (1.3, 4.0) | 2.3 (0.3, 4.3) |
| Between-group difference, mean (95% CI) mmHg^a^ | −0.4 (−2.7, 2.0); p = 0.739 | |
| Relative IOP reduction, mean (95% CI) % | 14.4 (6.5, 22.3) | 13.2 (0.2, 26.2) |
| Reduction of 20% or more, n (%) | 5 (33.3) | 4 (28.6) |
| Reduction of 30% or more, n (%) | 2 (13.3) | 2 (14.3) |
| IOP 15 mmHg or below, n (%) | 9 (60.0) | 11 (78.6) |
| IOP 12 mmHg or below, n (%) | 4 (26.7) | 9 (64.3) |
| Baseline medications, mean (SD) n | 1.1 (1.2) | 2.2 (1.7) |
| Month-12 medications, mean (SD) n | 1.1 (1.2) | 1.5 (1.6) |
| Medication change, mean (95% CI) n | 0.0 (−0.6, 0.6) | −0.7 (−1.2, −0.2) |

CI confidence interval, IOP intraocular pressure, MD mean deviation, SD standard deviation.

Severity staged from baseline visual field MD using the Hodapp–Parrish–Anderson convention. Baseline visual field data were available for 146 of 260 eyes; 139 of these (83 one-spacer, 56 two-spacer) had an analyzable month-12 IOP after censoring and provide the denominators shown. IOP and medication values recorded after a secondary IOP-lowering procedure were censored. Absolute reduction is the mean of per-eye changes from baseline; percentage reduction is the mean of per-eye percentage changes, each referenced to that eye’s own baseline (see Methods). Threshold attainment reflects both baseline IOP and the magnitude of reduction; in the severe stratum, 6 of 14 two-spacer eyes versus 1 of 15 one-spacer eyes were already at or below 12 mmHg at baseline.

a. Two spacers minus one spacer in mean absolute IOP reduction, by unadjusted Welch t-test; positive values favor two spacers. Severity-by-spacer interaction p = 0.962 (change-score model) and p = 0.381 (ANCOVA for month-12 IOP adjusted for baseline IOP).
